# Prognosis and personalized *in-silico* prediction of treatment efficacy in cardiovascular and chronic kidney disease: a proof-of-concept study

**DOI:** 10.1101/2023.08.01.23293501

**Authors:** Mayra Alejandra Jaimes Campos, Iván Andújar, Felix Keller, Gert Mayer, Peter Rossing, Jan A Staessen, Christian Delles, Joachim Beige, Griet Glorieux, Andrew L Clark, William Mullen, Joost P Schanstra, Antonia Vlahou, Kasper Rossing, Karlheinz Peter, Alberto Ortiz, Archie Campbell, Frederik Persson, Agnieszka Latosinska, Harald Mischak, Justyna Siwy, Joachim Jankowski

## Abstract

**Background:** kidney and cardiovascular diseases are responsible for a large fraction of population morbidity and mortality. Early, targeted, personalized intervention represents the ideal approach to cope with this challenge. Proteomic/peptidomic changes are largely responsible for onset and progression of these diseases and should hold information about optimal means for treatment and prevention.

**Methods:** we investigated prediction of renal or cardiovascular events using previously defined urinary peptidomic classifiers CKD273, HF2 and CAD160 in a cohort of 5585 subjects in a retrospective study.

**Results:** we demonstrate highly significant prediction of events with HR of 2.59, 1.71, and 4.12 for HF, CAD and CKD respectively. We applied in silico treatment, implementing on each patient urinary profile, changes onto the classifiers corresponding to exactly defined peptide abundance changes following commonly used interventions (MRA, SGLT2i, DPP4i, ARB, GLP1RA, olive oil and exercise), as defined in previous studies. Applying the proteomic classifiers after in silico treatment indicated individual benefits of specific interventions on a personalized level.

**Conclusions:** the *in-silico* evaluation may provide information on the future impact of specific drugs and intervention on endpoints, opening the door to a precision medicine approach. Investigation of the extent of the benefit of this approach in a prospective clinical trial is warranted.

## 1. Introduction

Cardiovascular diseases, including coronary artery disease (CAD) and heart failure (HF), along with chronic kidney disease (CKD), are leading causes of morbidity and mortality worldwide [1,2]. These conditions place a significant burden on affected individuals and healthcare systems globally. Efforts to reduce known cardiovascular and kidney risk factors, such as hypertension, high cholesterol levels, sedentary lifestyle, diabetes, obesity, and smoking, help prevent disease progression in some patients [2,3]. Advances in medical care and novel treatments have improved the prognosis of individuals affected with these chronic diseases [1,4,5]. Despite this progress, the factors associated with disease progression in individual patients are poorly understood. While traditional clinical risk factors and underlying molecular mechanisms explain a significant part of attributable risk [6,7], their predictive power for future cardiovascular or kidney events is limited or has not been evaluated and, in certain cases, may not be readily applicable in clinical setting [7–11].

Furthermore, CAD, HF and CKD require complex treatment regimen comprising multiple drugs combinations. Randomised trials demonstrate the value of different individual treatments in preventing future cardiac or kidney events, reducing mortality, and man-aging symptoms [12–16]. However, the benefit of a treatment is only detected in some patients and, a substantial number of individuals still progress to terminal organ failure, despite the treatment. Commonly recommended treatments include lifestyle interventions including dietary changes, antiplatelet therapy, β-blockers, angiotensin-converting enzyme inhibitors (ACEI), angiotensin receptor blockers (ARB), mineralocorticoid receptor antagonists (MRAs), glucagon-like peptide-1 receptor agonists (GLP1 RAs), dipeptidyl peptidase-4 inhibitors (DPP4i), and sodium-glucose co-transporter 2 inhibitors (SGLT2i) [17,18]. However, while these drugs demonstrably have an impact on notional targets, such as reduction of blood pressure or blood glucose, the targets are often surrogates for the real reason to treat -that is, preventing (or delaying) end-organ damage. That is more difficult to assess and needs a much longer time scale than days or weeks. There are currently no methods to predict treatment success in individuals or to give guidance on the optimal therapy for an individual patient.

Recent advances in biomarker research have contributed to the development of predictive classifiers that are more accurate markers of the progression towards adverse outcomes, including severe disease or mortality [7,19,20]. Multidimensional urinary peptides profiles seem to be particularly helpful in predicting outcome at early stage and can show the effect of treatment in different chronic diseases at molecular level [7,21–24].

To the best of our knowledge, no study has yet investigated the potential ability of a bi-omarker-based information to predict the potential impact of different interventions in decreasing the risk of events (critical progression or death) from cardiovascular or kidney diseases on a personalized level. The objective of this study was 1) to assess the efficacy of three previously developed urinary peptide-based classifiers as biomarkers for predicting CAD, HF or CKD events, and 2) to investigate in silico the individual impact of prophylactic or therapeutic interventions with specific active agents, with the hypothesis that the treatment that shows the most pronounced effect in silico, should present the optimal personalized therapeutic strategy.

## 2. Materials and Methods

### Study participants and study design

This study included 5585 datasets from previous studies: PRIORITY, DIRECT, FLE-MENGHO, CACTI, CardioRen, CAD prediction, Generation Scotland, HOMAGE, SUNmacro, and UZ-Gent. Detailed information on the designs and the methods used in these studies are available in previous publications [11,25–37]. Inclusion criteria were availability of estimated glomerular filtration rate (eGFR, calculated using the CKD Epidemiology Collaboration (CKD-EPI) formula), information on cardiovascular events, and availability of follow-up information. The endpoints were defined as follows: for coronary artery disease, the event was defined as non-fatal and fatal acute myocardial infarction. A heart failure event was defined as hospitalisation or death from heart failure. For CKD, an event was defined as a decline of ≥40% in eGFR values during follow-up, and the date when this decline was observed was considered as the duration of follow-up. Only one (the first) endpoint per patient was allowed: if an endpoint was reached, further endpoints was censored.

All individuals with urine samples at the baseline visit were included in the analysis. Several covariables including body mass index, age, sex, blood pressure and eGFR, were determined at the time of the baseline assessment. The median follow-up was 3.74 ±3.36 years. The study was conducted according to the guidelines of the Declaration of Helsinki and all datasets were fully anonymized. This study was approved by the ethics committee of the Hannover Medical School Germany under the reference number 3116-2016.

### Peptide-based classifiers and prediction of events

The classifier CKD273 was used for prediction of CKD events and impact of treatments [20]. The predictive capacity of, and impact of treatments on, the classifiers, HF2 and CAD-160-marker, were assessed for HF and CAD, respectively [30,38]. The scores for each classifier were calculated using a support vector machine (SVM) algorithm, integrated into the MosaCluster software [39]. All statistical tests were performed in R statistical software (R version 4.1.0, R Foundation for Statistical Computing, Vienna, Austria). The Kaplan-Meier estimator was applied to assess the association of longitudinal survival with each classifier. Corresponding hazard ratios (HR) were estimated using Cox regression models and log-rank tests were used to assess the hypothesis of no group differences in hazard functions. All survival analyses were carried out using the R package “survival”.

### *In-silico* impact of treatments

To assess the impact of various treatments on the classifiers for CAD, HF and CKD, the impact on urinary peptidomic profiles from five different drug-based interventions (MRA, SGLT2i, GLP1RA, DPP4i and ARB), one dietary intervention (olive oil) or from exercise was applied. These data were generated in previous studies that were either published, submitted for publication studies [24,40– 42], or unpublished (exercise). Briefly, the effect of the interventions on the urinary peptidomic profiles was assessed and the fold change values (as a result of the intervention) were determined. To predict the impact of treatment, these fold changes were then used to multiply the intensities of the respective peptides in each patient, and the predictor (CKD273, HF2, CAD-160-marker) scores were re-calculated. A decrease in the classifier score is indicative of a positive impact of the treatment on the outcome, as depicted in **Figure 1**. The results were visually represented using heatmaps generated with the R package “ComplexHeatmap”.

**Figure 1.**
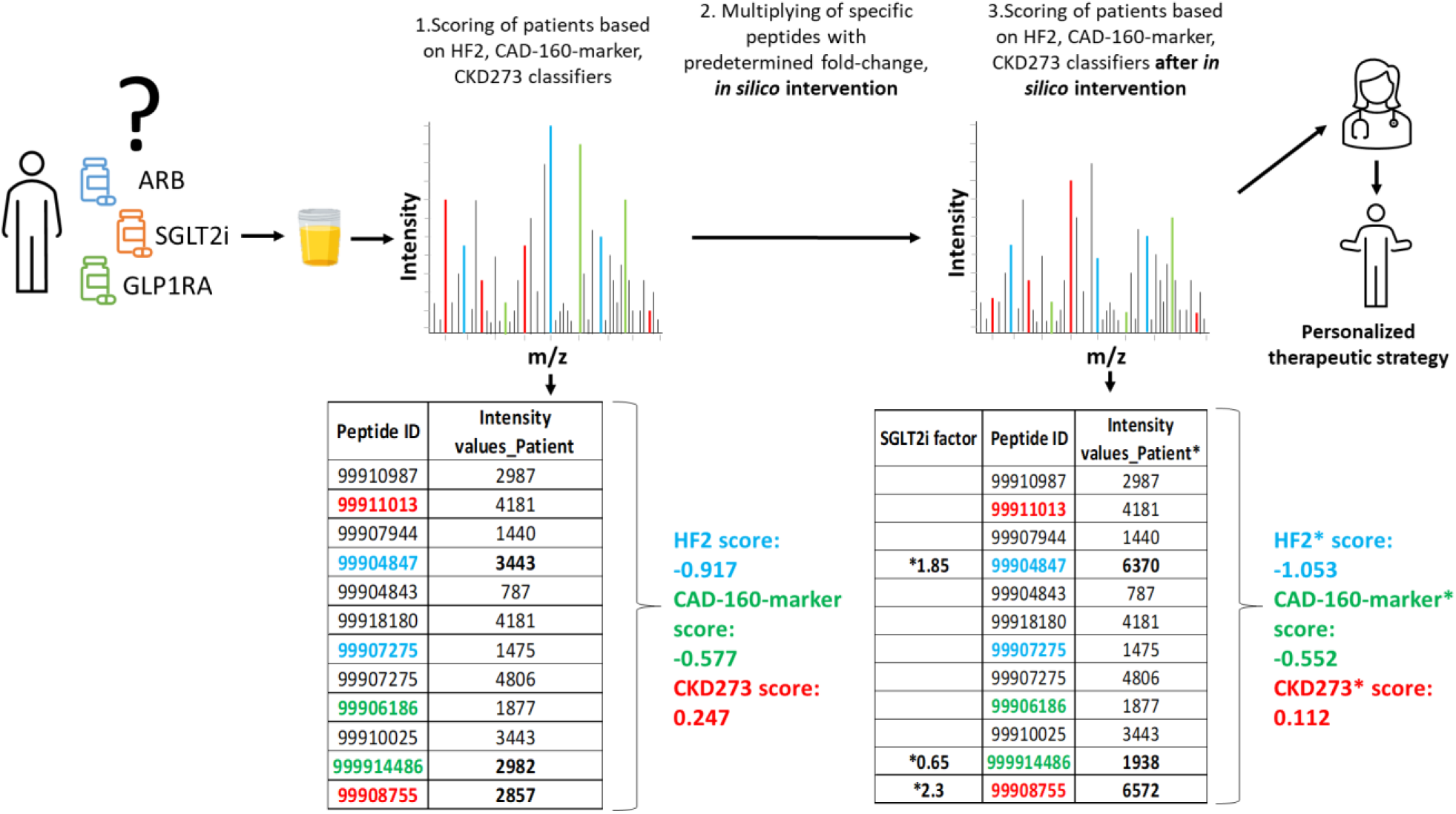
Schematic depiction of the study design. The relative abundance of 5071 se-quenced urinary peptides was investigated using CE-MS. Data on some selected peptides (ID) for 1 subject are shown. Several of these peptides were previously identified as being associated with the respective pathophysiology and combined into classifiers, CKD273, CAD160, and HF2. Some of these peptides are labelled with the respective color. In the first step, the patient receives a score for progression to event using the predefined urinary classifiers. Of the peptides shown, 3, labelled in bold, were found to be affected by SGLT2i treatment. In the second step, abundance of these 3 peptides is adjusted based on the observed fold change as a result of the treatment (“in silico treatment”). The classifier score is then re-calculated (labelled *) and the result is compared to the initial scoring; decrease in the scoring indicates benefit of the treatment. In this example, relevant impact of the SGLT2i treatment on CKD and HF event is predicted, but not impact on CAD.

## 3. Results

### 3.1 Clinical characteristics of population

A total of 5585 datasets was extracted from the database. Baseline characteristics are shown in **Table 1**

**Table 1.**
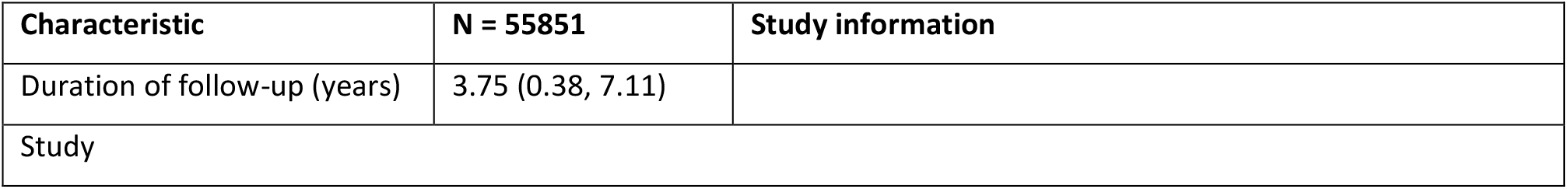
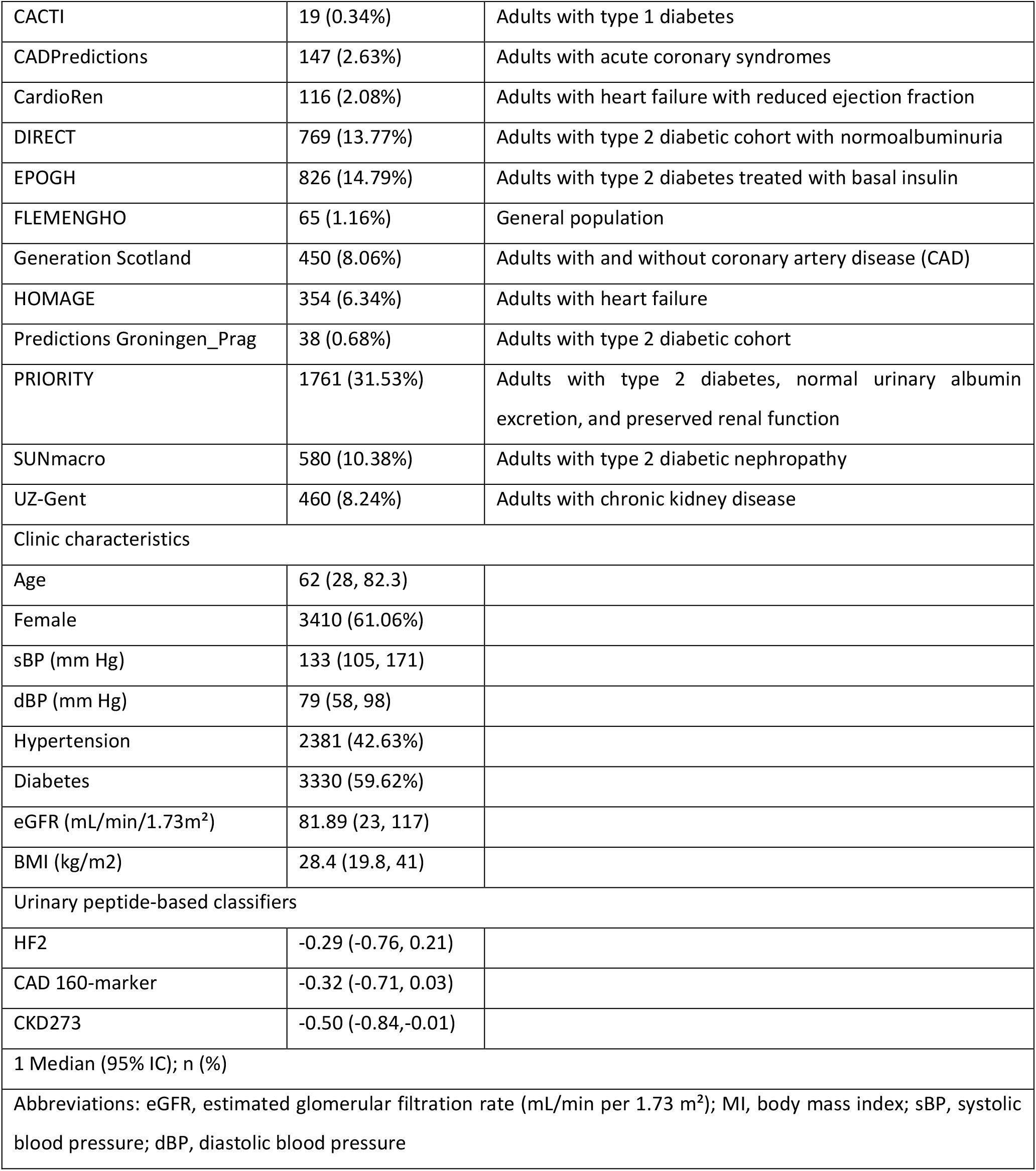
Studies information and baseline characteristics of study participants.

### 3.2 Peptide-based classifiers and prediction of events

The association between the classifiers and the risk of cardiovascular/kidney events is detailed in **Table 2**. Individuals were divided into quintiles with different relative risk according to their classifier scores (**Table S1**). The event rates for the outcome of cardio-vascular/kidney events varied across the five score subgroups. There was a stepwise in-crease in the risk of an adverse event with each quintile, that is, individuals with higher classifier scores, as represented in the 5th quintile, had higher rates of the primary out-come compared to individuals in the lower quintile of the classifier (1st quintile) (**Figure 2**).

**Table 2.**
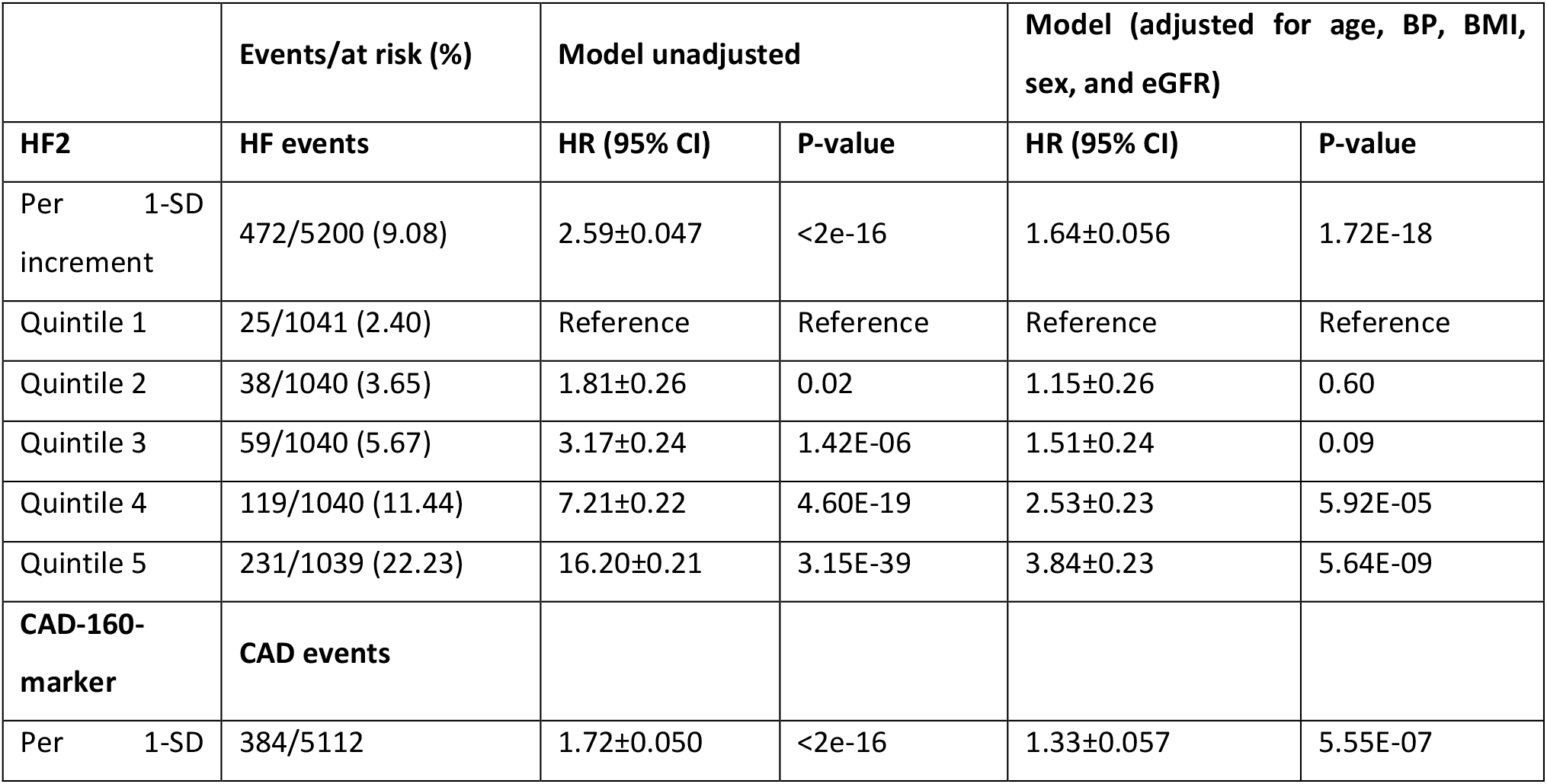
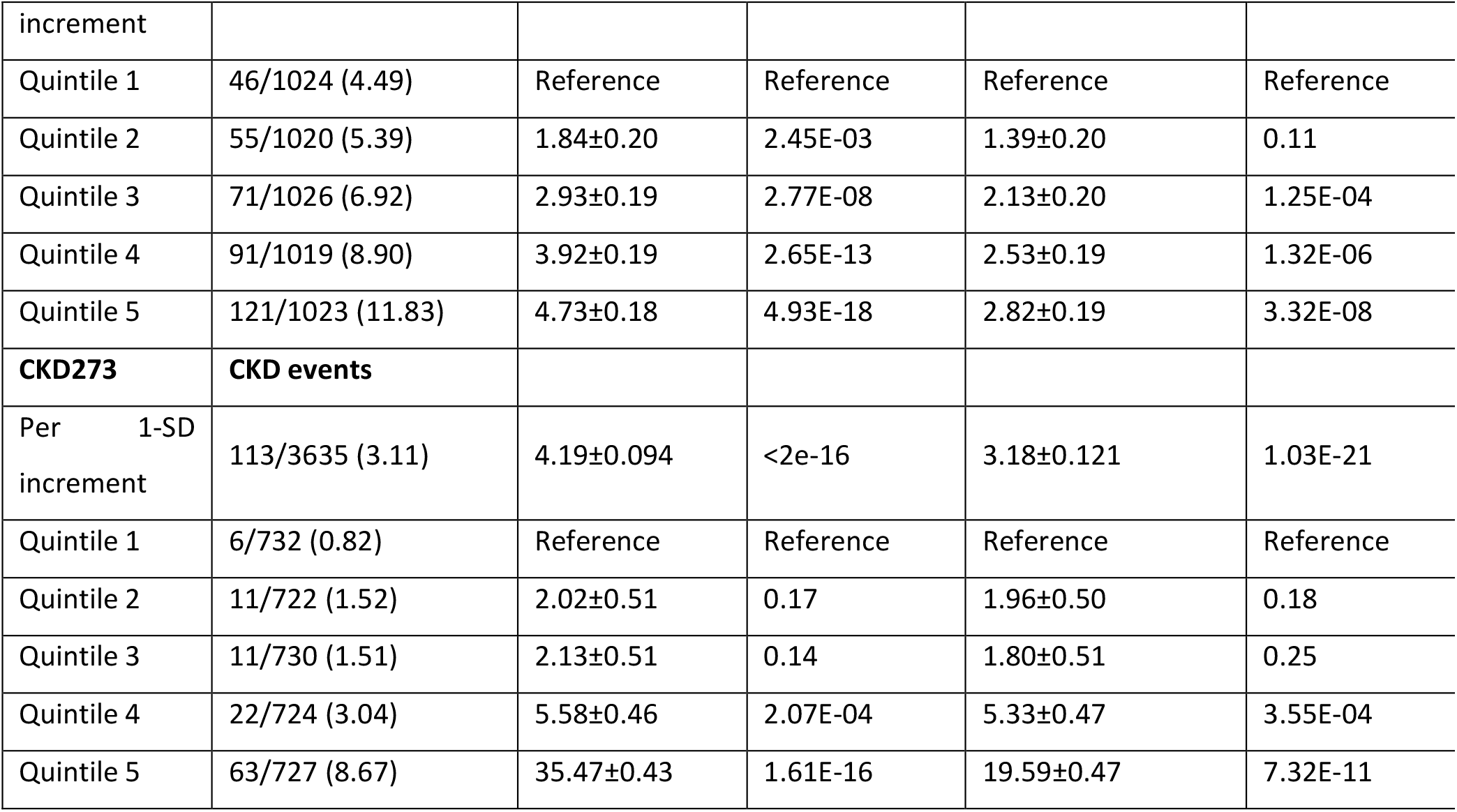
Risk of HF events, CAD events and CKD outcomes by baseline urinary peptidomics classifier.

**Figure 2.**
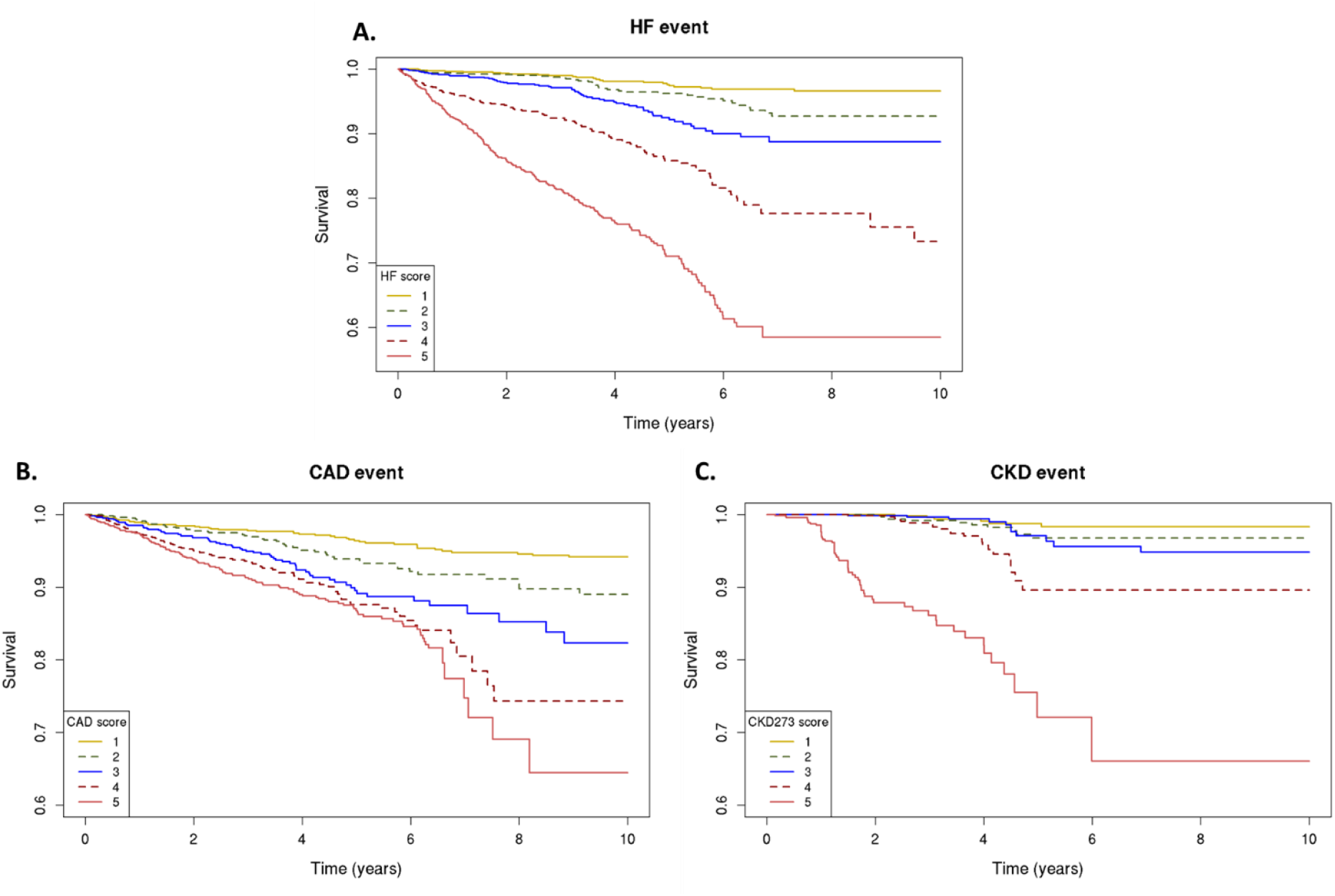
Urinary peptidomics classifiers and primary outcomes. Kaplan-Meier curves for the primary outcome; classifier scores from lowest (1) to highest (5) quintile, for risk of heart failure events as assessed by HF2 (A), coronary artery disease events as assessed by CAD160 (B), and chronic kidney disease progression as assessed by CKD273 (C).

### 3.3 Personalized *in-silico* prediction of treatment efficacy

Having established a highly significant association between the classifiers scores and outcomes, we investigated whether the *in-silico* treatment effect (e.g., adjustment of the peptide intensities based on the treatment response) as described in the Methods section, had an impact on the classifiers. The *in-silico* treatment, had a significant impact on the classifiers, as shown in **Figures 3, 4** and **5** and also given in **Table S1**. The heatmap representation of the sorted scores before the *in-silico* treatment revealed an alignment of HF events, CAD events, and CKD progression with higher scores (**Figure 3. 4 and 5**). This observation reinforces the predictive capability of the scores and their association with HF and CAD events and CKD progression.

**Figure 3.**
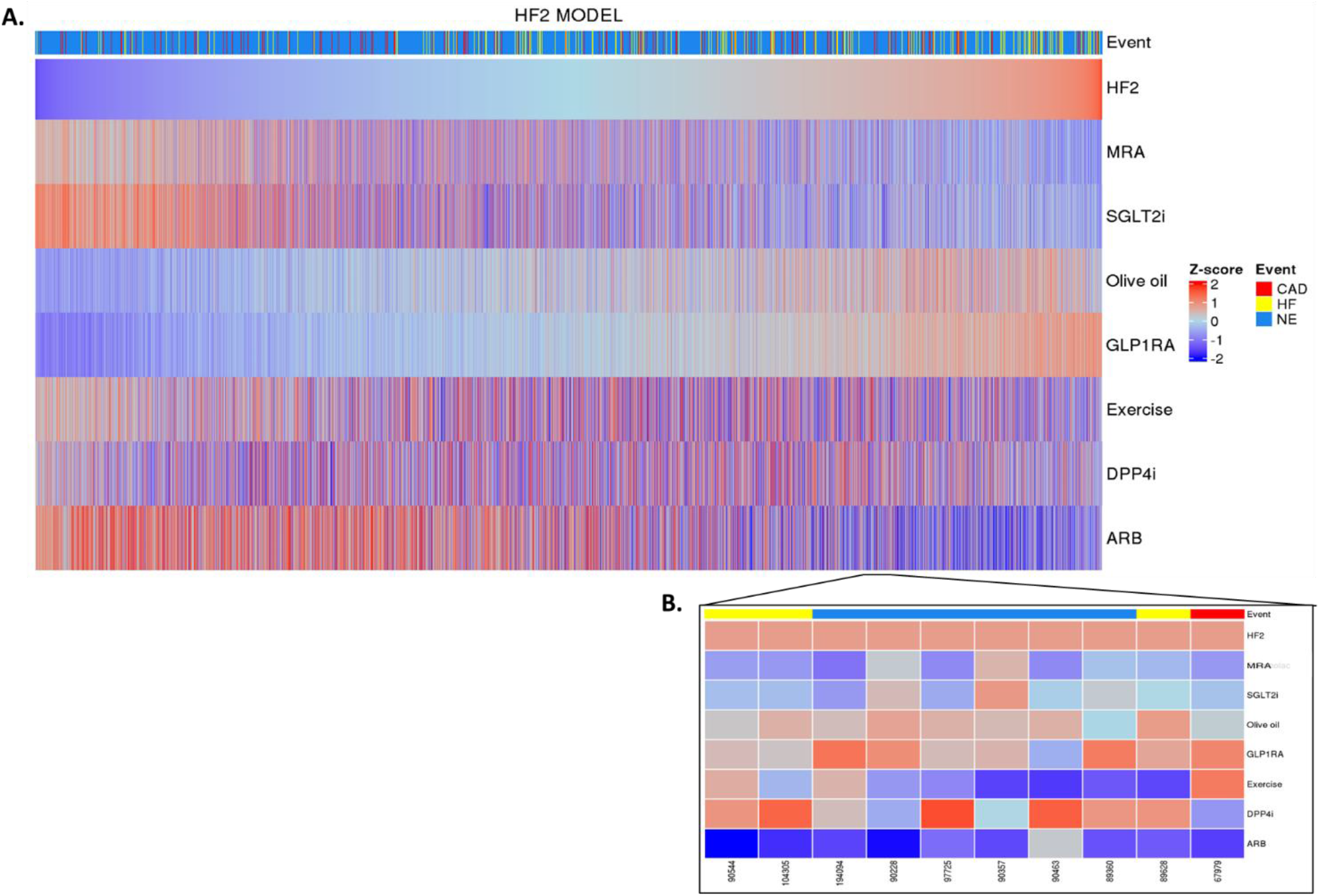
HF2 classifier treatment responses. HF2 scores were z-scaled across samples for visualisation. Heatmap HF2 classifier treatment responses of 5585 patients (A). The top of the heatmap shows event information. Samples (columns) were ordered based on HF2 score prior to in silico treatment; from lower scores (left) to higher scores (right). Zoomed heatmap shows the treatment response of HF2 in 10 patients (B). Patients who were already receiving one of the treatments at the beginning of the study are depicted in grey.

**Figure 4.**
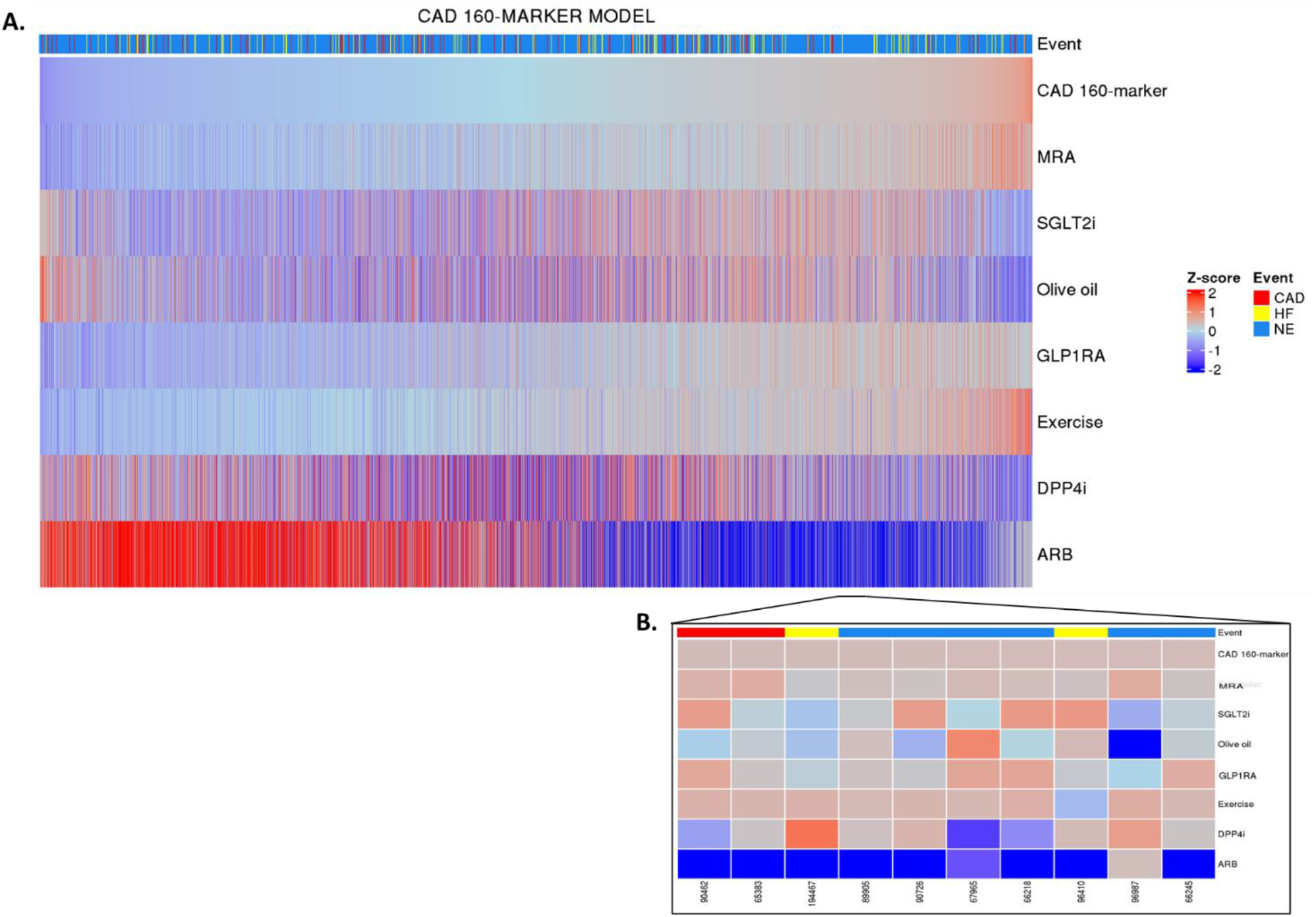
CAD-160-marker classifier treatment responses. CAD-160-marker scores were z-scaled across samples for visualisation. Heatmap CAD-160-marker classifier treatment responses of 5585 patients (A). The top of the heatmap shows event information. Samples (columns) were ordered based on CAD-160-marker score prior to in silico treatment; from lower scores (left) to higher scores (right). Zoomed heatmap shows the treatment response of the CAD-160-marker classifier in 10 patients (B). Patients who were already receiving one of the treatments at the beginning of the study are depicted in grey.

**Figure 5.**
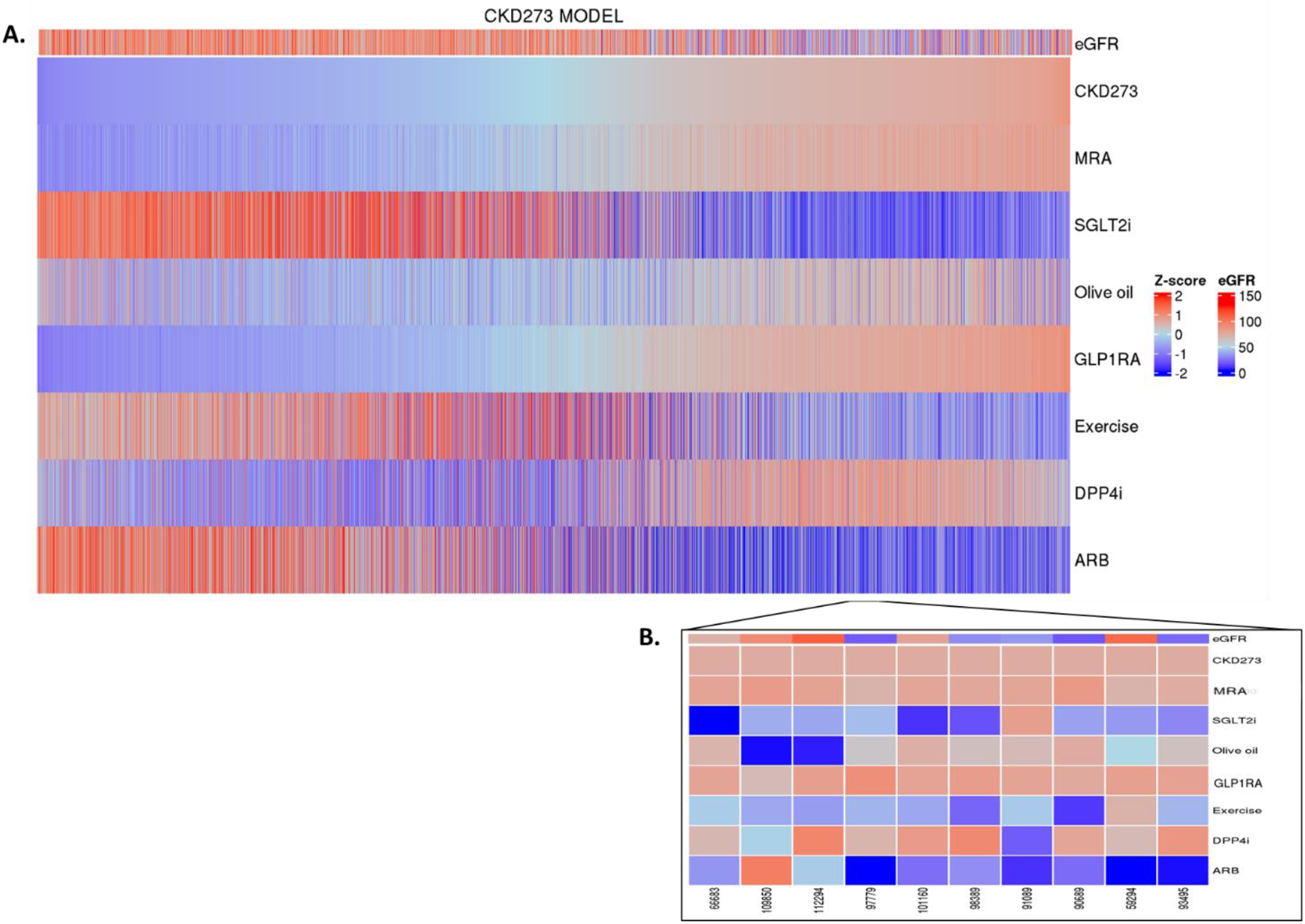
CKD273 classifier treatment responses. CKD273 scores were z-scaled across samples for visualisation. Heatmap CKD273 classifier treatment responses of 5585 pa-tients (A) The top of the heatmap shows baseline eGFR values information. Samples (columns) were ordered based on CKD273 score prior to in silico treatment; from lower scores (left) to higher scores (right). Zoomed heatmap shows the treatment response of the CKD273 classifier in 10 patients (B). Patients who were already receiving one of the treatments at the beginning of the study are depicted in grey.

After the *in-silico* treatment, there was a positive effect of MRA, SGLT2i, and ARB treatments on the HF2 classifier in individuals with higher scores, suggesting a potential beneficial impact of treatment especially in those at higher baseline risk (and likely more advanced disease (**Figure 3A**). Olive oil and GLP1R agonist treatment showed a positive impact mostly in individuals at low risk of HF events. DPP4i and exercise had inconsistent effects across different scores, making its impact on patients with high risk of HF events less evident. Predictions showed individual differences in the treatment impact (**Figure 3B**).

Regarding the CAD-160-marker classifier, distinctly different treatment responses were observed when comparing the CAD-160-marker score-high and CAD-160-marker score-low groups (**Figure 4A**). Among individuals with higher scores, olive oil, DPP4i, and especially ARB treatments were predicted to present positive impacts, with ARB treatment being notably effective for patients at high risk of CAD events. Nonetheless, individual predictions displayed unique differences, emphasizing the personalized nature of pre-diction of treatment response (**Figure 4B**).

In the context of CKD, no major impact was observed for spironolactone and for GLP1RA. In patients with high CKD273 scoring, many of these having eGFR values less than 60 mL/min per 1.73 m^2^, SGLT2i, olive oil, exercise, and ARB treatments exhibited treatment responses, with SGLT2i and ARB treatments showing a more pronounced impact. In contrast, olive oil treatment seemed to have a positive impact mostly in patients with lower scores (**Figure 5A and B**). The impact of DPP4i varies among patients, and in certain cases, it demonstrates a positive effect in advanced stages of the disease.

## 4. Discussion

The identification of biomarkers that aid physicians in decision-making and treatment planning for patients with cardiovascular or kidney disease will serve a major clinical need. Early diagnosis of cardiovascular and CKD is challenging, as patients may remain asymptomatic in the early stages, leading to late-stage clinical presentations and diagnosis/detection. Additionally, considering the significant interindividual variability in response to different treatments, uncertainty remains about whether the development of an event can be avoided in a patient during drug prescription. While a substantial number of studies demonstrated potential value of biomarkers in predicting disease progression and renal as well as cardiovascular events [7-11], these studies typically did not investigate the in fact more relevant topic (from the patient point-of-view): prediction of optimal intervention. Prediction of drug response on a population-based level was proposed by the group from Heerspink [43], but not on an individual level. Therefore, the crucial need for non-invasive biomarkers for early disease detection and to understand the impact of different treatments becomes evident, enabling timely treatment and prevention of chronic progression.

Multiple drugs are available that impact on risk factors like elevated blood pressure, blood glucose or cholesterol. Normalization of these parameters can generally be easily and rapidly assessed. However, the question if normalization of these parameters has a beneficial impact on target organ damage on an individual level cannot be easily answered and would require long-term follow-up, not compatible with clinical practice. What is needed would be an approach to assess, ideally even predict the impact of the drug on outcome, on the target organ damage. In this study, we assessed three established urinary peptide-based classifiers: HF2, CAD-160-marker, and CKD273 [20,30,38], designed to predict the risk of major complications or mortality in individuals at high risk of, or already suffering from chronic cardiovascular or kidney disease conditions and we investigated the potential impact of different interventions on reducing the occurrence of these events.

The study presented here has two main results. The first result is the demonstration of the prognostic value of the three applied urinary peptide classifiers. This result further confirms the previous reports [20,30,38] in large cohorts. While such prognosis is valuable in guiding treatment and management, it obviously lacks specific guidance on the treatment. This fact leads to the second main result: applying the previously established impact of specific treatment on the urinary peptides allows implementation of “*in-silico* treatment”, which may be used to guide, personalized intervention, based on the predicted response. Using this *in-silico* approach, we achieved individualised prediction of the effects of seven different treatment approaches based on the urinary peptide-based classifiers. These findings offer a novel approach towards personalised treatment strategies and risk management for patients at risk of cardiovascular or kidney diseases, based on the predicted molecular impact of the specific treatment.

Previous studies have already demonstrated the predictive performance of the HF2 model, the CAD-160-marker model and the CKD273 model in different populations for the respective clinical conditions [20,30,38]. In our larger population, we observed effective risk stratification based on model scores, successfully identifying patients at higher risk of cardiovascular/kidney events. Specifically, individuals in the lower score group exhibited a reduced risk of HF, CAD and CKD events compared to those in the higher score group.

Regarding the individualised prediction of treatment impact, we observed significant effects with interventions such as SGLT2i, ARB, MRA, DPP4i, and lifestyle interventions. The overall observations are consistent with the results from previous intervention studies. Specifically, it appears that a benefit of ARB in CKD is most prominent in subjects with the highest risk, likely in late-stage disease, which is in agreement with failure to demonstrate a significant benefit of ARB at early-stage disease [44]. We also detect more pronounced benefit of SGLT2i, particularly in subjects with high risk of HF and CKD, but to a much lesser degree in the context of CAD, which is in very good agreement with the respective intervention studies [45–48]. Impact in the context of CAD appears most prominent for ARB in subjects with increased risk, which was also observed in the intervention trials [49,50]. MRA is a recommended treatment in HF for individuals with reduced and preserved ejection fraction. Notably, our findings revealed a benefit effect of MRA in HF among individuals at the highest risk but not as clear in CAD or CKD, in line also with the results of the PRIORITY trial [11]. Our results are consistent with previous clinical trials, supporting the potential efficacy of MRA improve HF outcomes [51]. However, the impact of MRA in CAD or CKD is not clearly demonstrated. In CKD, MRA showed an early effect on renal function changes but did not have longer-term effects [52].

Several studies have investigated the potential beneficial effects of GLP1RA and DPP4i in cardiovascular and kidney disease [53–56]. Some clinical studies have suggested that these drugs may have a beneficial impact on the progression of these conditions, but data from clinical trials remain somewhat controversial [57]. In our study, we observed a positive impact in individuals with higher risk of CAD. However, further trials with appropriate power and design are necessary. Larger and well-controlled clinical trials will provide a clearer understanding of the potential benefits of these drugs.

Lifestyle modification is generally recommended for the management of cardiovascular and kidney diseases. However, when it comes to CKD and CAD, physical activity recommendations should be carefully considered depending on the patient’s condition due to the potential risk of impairing kidney function and increasing proteinuria, or triggering cardiovascular events during exercise [58,59]. As for olive oil in the diet, some evidence suggested that this intervention may have a beneficial impact in preventing cardiovascular or kidney events [60,61]. Specifically, our observations revealed that the impact of the lifestyle intervention seems to have an individual pattern, with a positive impact in patients at both low and high risk. Regarding exercise, we observe a positive impact in individuals at lower risk of CAD events and in some individuals at high risk. Meanwhile, it seems to have a preventive effect on HF and CKD events in individuals at high risk, which is consistent with findings from previous studies [62,63].

The study also has shortcomings and the results should be interpreted with caution. First, this is a retrospective study based on data collected in the context of multiple different previous studies. However, the large number of subjects included is expected to counteract potential bias introduced by some of the specific previous cohorts. Further, the results observed are fully in line with previous observations, further supporting the validity. Second, the impact of treatment on the urinary proteome by the different means of intervention is not fully comparable, number of subjects in these previous studies, demographic characteristic, and duration of intervention differed. To counteract these issues, data were normalized to Z-scores to prevent dominance of one specific intervention. Given the results, that some types of intervention are predicted to be specifically beneficial in certain situations (e.g., SGLT2 inhibition indicating benefit in most patients with a high risk of HF and CKD events, but not in subjects with high risk of CAD events), and that this observation is in very good agreement with the observations in the intervention trials reported, further support the validity of the approach. At the same time, inter-individual variability is observed which highlights the personalized aspect of the presented *in-silico* predictor, to be further tested in the context of a prospective clinical intervention trial. Along the same lines, we have not formally demonstrated that the prediction of the best suited intervention does in fact give a significant benefit to patients with respect to preventing experiencing any of the patient-relevant endpoints. Such a benefit can only be demonstrated in a prospective trial. However, based on the data available, we feel that using this approach may well be justified in a situation when guidance on the ideal intervention is missing. In addition, we are currently in the process of initiating the prospective trial that would demonstrate a significant benefit.

In conclusion, the *in-silico* evaluation of the impact of different drugs in an individual`s urinary peptidomics signature may provide information on the future impact of specific drugs on hard endpoints in this individual, opening the door to a precision medicine approach to select the optimal treatment for individuals with or at risk of CAD, HF or CKD progression. The performance of this *in-silico* test should be validated in a prospective clinical trial.

## Data Availability

All data produced in the present work are contained in the manuscript

## Supplementary Materials

Table S1: Classifier score before and after the *in-silico* treatment.

## Author Contributions

Conceptualization, MAJC, HM, JS, AV, JPS and AL; methodology, MAJC, AL, GM, PR, JAS, CD, AC, JB, GG, ALC, WM, KR, KP, AO, JJ, FP and JS; formal analysis, MAJC, IA, AL, FK and JS; writing—original draft preparation, MAJC; visualization, MAJC. All authors have read and agreed to the published version of the manuscript.

## Institutional Review Board Statement

The study was conducted in accordance with the Declara-tion of Helsinki and approved by the ethics committee of the Hannover Medical School Germany under the reference number 3116-2016. Informed Consent Statement: Informed consent was obtained from all subjects involved in the study.

## References

1. Ezzati, M.; Obermeyer, Z.; Tzoulaki, I.; Mayosi, B.M.; Elliott, P.; Leon, D.A. Contributions of Risk Factors and Medical Care to Cardiovascular Mortality Trends. Nat. Rev. Cardiol. 2015, 12, 508–530; doi:10.1038/nrcardio.2015.82.

2. Webster, A.C.; Nagler, E. V.; Morton, R.L.; Masson, P. Chronic Kidney Disease. Lancet 2017, 389, 1238–1252; doi:10.1016/S0140-6736(16)32064-5.

3. Bays, H.E.; Taub, P.R.; Epstein, E.; Michos, E.D.; Ferraro, R.A.; Bailey, A.L.; Kelli, H.M.; Ferdinand, K.C.; Echols, M.R.; Wein-traub, H.; et al. Ten Things to Know about Ten Cardiovascular Disease Risk Factors. Am. J. Prev. Cardiol. 2021, 5, 100149; doi:https://doi.org/10.1016/j.ajpc.2021.100149.

4. Tzoulaki, I.; Elliott, P.; Kontis, V.; Ezzati, M. Worldwide Exposures to Cardiovascular Risk Factors and Associated Health Ef-fects: Current Knowledge and Data Gaps. Circulation 2016, 133, 2314–2333; doi:10.1161/CIRCULATIONAHA.115.008718.

5. Chen TK, Knicely DH G.M. Chronic Kidney Disease Diagnosis and Management: A Review. 2019, 322, 1294–1304; doi:10.1001/jama.2019.14745.

6. Sinha, A.; Ning, H.; Carnethon, M.R.; Allen, N.B.; Wilkins, J.T.; Lloyd-Jones, D.M.; Khan, S.S. Race- and Sex-Specific Population Attributable Fractions of Incident Heart Failure. Circ. Hear. Fail. 2021, 14, e008113; doi:10.1161/CIRCHEARTFAILURE.120.008113.

7. Pontillo, C.; Zhang, Z.-Y.; Schanstra, J.P.; Jacobs, L.; Zürbig, P.; Thijs, L.; Ramírez-Torres, A.; Heerspink, H.J.L.; Lindhardt, M.; Klein, R.; et al. Prediction of Chronic Kidney Disease Stage 3 by CKD273, a Urinary Proteomic Biomarker. Kidney Int. reports 2017, 2, 1066–1075; doi:10.1016/j.ekir.2017.06.004.

8. Vasan, R.S. Biomarkers of Cardiovascular Disease: Molecular Basis and Practical Considerations. Circulation 2006, 113, 2335–2362; doi:10.1161/CIRCULATIONAHA.104.482570.

9. Hense, H.-W. Observations, Predictions and Decisions—Assessing Cardiovascular Risk Assessment. Int. J. Epidemiol. 2004, 33, 235–239; doi:10.1093/ije/dyh118.

10. Musunuru, K.; Hershberger, R.E.; Day, S.M.; Klinedinst, N.J.; Landstrom, A.P.; Parikh, V.N.; Prakash, S.; Semsarian, C.; Sturm, A.C. Genetic Testing for Inherited Cardiovascular Diseases: A Scientific Statement From the American Heart Associa-tion. Circ. Genomic Precis. Med. 2020, 13, E000067; doi:10.1161/HCG.0000000000000067.

11. Tofte, N.; Lindhardt, M.; Adamova, K.; Bakker, S.J.L.; Beige, J.; Beulens, J.W.J.; Birkenfeld, A.L.; Currie, G.; Delles, C.; Dimos, I.; et al. Early Detection of Diabetic Kidney Disease by Urinary Proteomics and Subsequent Intervention with Spironolactone to Delay Progression (PRIORITY): A Prospective Observational Study and Embedded Randomised Placebo-Controlled Trial. Lancet Diabetes Endocrinol. 2020, 8, 301–312; doi:10.1016/S2213-8587(20)30026-7.

12. Ziff, O.J.; Samra, M.; Howard, J.P.; Bromage, D.I.; Ruschitzka, F.; Francis, D.P.; Kotecha, D. Beta-Blocker Efficacy across Dif-ferent Cardiovascular Indications: An Umbrella Review and Meta-Analytic Assessment. BMC Med. 2020, 18, 103; doi:10.1186/s12916-020-01564-3.

13. Andrikou, E.; Tsioufis, C.; Andrikou, I.; Leontsinis, I.; Tousoulis, D.; Papanas, N. GLP-1 Receptor Agonists and Cardiovascular Outcome Trials: An Update. Hell. J. Cardiol. 2019, 60, 347–351; doi:https://doi.org/10.1016/j.hjc.2018.11.008.

14. Ong, H.T.; Ong, L.M.; Ho, J.J. Angiotensin-Converting Enzyme Inhibitors (ACEIs) and Angiotensin-Receptor Blockers (ARBs) in Patients at High Risk of Cardiovascular Events: A Meta-Analysis of 10 Randomised Placebo-Controlled Trials. ISRN Cardiol. 2013, 2013, 478597; doi:10.1155/2013/478597.

15. Patoulias, D.I.; Boulmpou, A.; Teperikidis, E.; Katsimardou, A.; Siskos, F.; Doumas, M.; Papadopoulos, C.E.; Vassilikos, V. Cardiovascular Efficacy and Safety of Dipeptidyl Peptidase-4 Inhibitors: A Meta-Analysis of Cardiovascular Outcome Trials. World J. Cardiol. 2021, 13, 585–592; doi:10.4330/wjc.v13.i10.585.

16. Usman, M.S.; Siddiqi, T.J.; Memon, M.M.; Khan, M.S.; Rawasia, W.F.; Talha Ayub, M.; Sreenivasan, J.; Golzar, Y. Sodium-Glucose Co-Transporter 2 Inhibitors and Cardiovascular Outcomes: A Systematic Review and Meta-Analysis. Eur. J. Prev. Cardiol. 2018 25, 495–502; doi:10.1177/2047487318755531.

17. Yang, Y.; Gao, J.; Qin, Z.; Lu, Y.; Xu, Y.; Guo, J.; Cui, X.; Zhang, J.; Tang, J. The Present Clinical Treatment and Future Emerging Interdisciplinary for Heart Failure: Where We Are and What We Can Do. Intensive Care Res. 2023, 3, 3–11; doi:10.1007/s44231-023-00029-4.

18. Shlipak, M.G.; Tummalapalli, S.L.; Boulware, L.E.; Grams, M.E.; Ix, J.H.; Jha, V.; Kengne, A.P.; Madero, M.; Mihaylova, B.; Tangri, N.; et al. The Case for Early Identification and Intervention of Chronic Kidney Disease: Conclusions from a Kidney Disease: Improving Global Outcomes (KDIGO) Controversies Conference. Kidney Int. 2021, 99, 34–47; doi:10.1016/j.kint.2020.10.012.

19. Wendt, R.; Thijs, L.; Kalbitz, S.; Mischak, H.; Siwy, J.; Raad, J.; Metzger, J.; Neuhaus, B.; Leyen, H. von der; Dudoignon, E.; et al. A Urinary Peptidomic Profile Predicts Outcome in SARS-CoV-2-Infected Patients. eClinicalMedicine 2021, 36, 6–7; doi:10.1016/j.eclinm.2021.100883.

20. Argilés, À.; Siwy, J.; Duranton, F.; Gayrard, N.; Dakna, M.; Lundin, U.; Osaba, L.; Delles, C.; Mourad, G.; Weinberger, K.M.; et al. CKD273, a New Proteomics Classifier Assessing CKD and Its Prognosis. PLoS One 2013, 8, e62837; doi:10.1371/journal.pone.0062837.

21. Zhang, Z.-Y.; Nkuipou-Kenfack, E.; Staessen, J.A. Urinary Peptidomic Biomarker for Personalized Prevention and Treatment of Diastolic Left Ventricular Dysfunction. Proteomics. Clin. Appl. 2019, 13, e1800174; doi:10.1002/prca.201800174.

22. Tofte, N.; Lindhardt, M.; Adamova, K.; Bakker, S.J.L.; Beige, J.; Beulens, J.W.J.; Birkenfeld, A.L.; Currie, G.; Delles, C.; Dimos, I.; et al. Early Detection of Diabetic Kidney Disease by Urinary Proteomics and Subsequent Intervention with Spironolactone to Delay Progression (PRIORITY): A Prospective Observational Study and Embedded Randomised Placebo-Controlled Trial. lancet. Diabetes Endocrinol. 2020, 8, 301–312; doi:10.1016/S2213-8587(20)30026-7.

23. Curovic, V.R.; Eickhoff, M.K.; Rönkkö, T.; Frimodt-Møller, M.; Hansen, T.W.; Mischak, H.; Rossing, P.; Ahluwalia, T.S.; Persson, F. Dapagliflozin Improves the Urinary Proteomic Kidney-Risk Classifier CKD273 in Type 2 Diabetes with Albuminuria: A Randomized Clinical Trial. Diabetes Care 2022, 45, 2662–2668; doi:10.2337/dc22-1157.

24. Siwy, J.; Klein, T.; Rosler, M.; von Eynatten, M. Urinary Proteomics as a Tool to Identify Kidney Responders to Dipeptidyl Peptidase-4 Inhibition: A Hypothesis-Generating Analysis from the MARLINA-T2D Trial. Proteomics. Clin. Appl. 2019, 13, e1800144; doi:10.1002/prca.201800144.

25. Lindhardt, M.; Persson, F.; Zürbig, P.; Stalmach, A.; Mischak, H.; de Zeeuw, D.; Lambers Heerspink, H.; Klein, R.; Orchard, T.; Porta, M.; et al. Urinary Proteomics Predict Onset of Microalbuminuria in Normoalbuminuric Type 2 Diabetic Patients, a Sub-Study of the DIRECT-Protect 2 Study. Nephrol. Dial. Transplant. 2017, 32, 1866–1873; doi:10.1093/ndt/gfw292.

26. Zhang, Z.; Staessen, J.A.; Thijs, L.; Gu, Y.; Liu, Y.; Jacobs, L.; Koeck, T.; Zürbig, P.; Mischak, H.; Kuznetsova, T. Left Ventricu-lar Diastolic Function in Relation to the Urinary Proteome: A Proof-of-Concept Study in a General Population. Int. J. Cardiol. 2014, 176, 158–165; doi:10.1016/j.ijcard.2014.07.014.

27. Kuznetsova, T.; Mischak, H.; Mullen, W.; Staessen, J.A. Urinary Proteome Analysis in Hypertensive Patients with Left Ven-tricular Diastolic Dysfunction. Eur. Heart J. 2012, 33, 2342–2350; doi:10.1093/eurheartj/ehs185.

28. Packham, D.K.; Wolfe, R.; Reutens, A.T.; Berl, T.; Heerspink, H.L.; Rohde, R.; Ivory, S.; Lewis, J.; Raz, I.; Wiegmann, T.B.; et al. Sulodexide Fails to Demonstrate Renoprotection in Overt Type 2 Diabetic Nephropathy. J. Am. Soc. Nephrol. 2012, 23, 123–130; doi:10.1681/ASN.2011040378.

29. Verbeke, F.; Siwy, J.; Van Biesen, W.; Mischak, H.; Pletinck, A.; Schepers, E.; Neirynck, N.; Magalhães, P.; Pejchinovski, M.; Pontillo, C.; et al. The Urinary Proteomics Classifier Chronic Kidney Disease 273 Predicts Cardiovascular Outcome in Patients with Chronic Kidney Disease. Nephrol. Dial. Transplant. Off. Publ. Eur. Dial. Transpl. Assoc. - Eur. Ren. Assoc. 2021, 36, 811–818; doi:10.1093/ndt/gfz242.

30. Snell-Bergeon, J.K.; Maahs, D.M.; Ogden, L.G.; Kinney, G.L.; Hokanson, J.E.; Schiffer, E.; Rewers, M.; Mischak, H. Evaluation of Urinary Biomarkers for Coronary Artery Disease, Diabetes, and Diabetic Kidney Disease. Diabetes Technol. Ther. 2009, 11, 1–9; doi:10.1089/dia.2008.0040.

31. Htun, N.M.; Magliano, D.J.; Zhang, Z.-Y.; Lyons, J.; Petit, T.; Nkuipou-Kenfack, E.; Ramirez-Torres, A.; von Zur Muhlen, C.; Maahs, D.; Schanstra, J.P.; et al. Prediction of Acute Coronary Syndromes by Urinary Proteome Analysis. PLoS One 2017, 12, e0172036; doi:10.1371/journal.pone.0172036.

32. Alkhalaf, A.; Zürbig, P.; Bakker, S.J.L.; Bilo, H.J.G.; Cerna, M.; Fischer, C.; Fuchs, S.; Janssen, B.; Medek, K.; Mischak, H.; et al. Multicentric Validation of Proteomic Biomarkers in Urine Specific for Diabetic Nephropathy. PLoS One 2010, 5, e13421; doi:10.1371/journal.pone.0013421.

33. Rossing, K.; Bosselmann, H.S.; Gustafsson, F.; Zhang, Z.-Y.; Gu, Y.-M.; Kuznetsova, T.; Nkuipou-Kenfack, E.; Mischak, H.; Staessen, J.A.; Koeck, T.; et al. Urinary Proteomics Pilot Study for Biomarker Discovery and Diagnosis in Heart Failure with Reduced Ejection Fraction. PLoS One 2016, 11, e0157167; doi:10.1371/journal.pone.0157167.

34. Smith, B.H.; Campbell, A.; Linksted, P.; Fitzpatrick, B.; Jackson, C.; Kerr, S.M.; Deary, I.J.; Macintyre, D.J.; Campbell, H.; McGilchrist, M.; et al. Cohort Profile: Generation Scotland: Scottish Family Health Study (GS:SFHS). The Study, Its Participants and Their Potential for Genetic Research on Health and Illness. Int. J. Epidemiol. 2013, 42, 689–700; doi:10.1093/ije/dys084.

35. He, T.; Mischak, M.; Clark, A.L.; Campbell, R.T.; Delles, C.; Díez, J.; Filippatos, G.; Mebazaa, A.; McMurray, J.J. V; González, A.; et al. Urinary Peptides in Heart Failure: A Link to Molecular Pathophysiology. Eur. J. Heart Fail. 2021, 23, 1875–1887; doi:10.1002/ejhf.2195.

36. Futter, J.E.; Cleland, J.G.F.; Clark, A.L. Body Mass Indices and Outcome in Patients with Chronic Heart Failure. Eur. J. Heart Fail. 2011, 13, 207–213; doi:10.1093/eurjhf/hfq218.

37. He, T.; Melgarejo, J.D.; Clark, A.L.; Yu, Y.-L.; Thijs, L.; Díez, J.; López, B.; González, A.; Cleland, J.G.; Schanstra, J.P.; et al. Serum and Urinary Biomarkers of Collagen Type-I Turnover Predict Prognosis in Patients with Heart Failure. Clin. Transl. Med. 2021, 11, e267. doi: 10.1002/ctm2.267

38. Wei, D.; Melgare, J.D.; Aelst, L. van; Vanassche, T.; Verhamme, P.; Janssens, S.; Peter, K.; Zhang, Z.-Y. Prediction of Coronary Artery Disease Using Urinary Proteomics. J. Hypertens. 2023, 41, e95; doi:10.1097/01.hjh.0000939684.90016.aa.

39. Theodorescu, D.; Schiffer, E.; Bauer, H.W.; Douwes, F.; Eichhorn, F.; Polley, R.; Schmidt, T.; Schöfer, W.; Zürbig, P.; Good, D.M.; et al. Discovery and Validation of Urinary Biomarkers for Prostate Cancer. Proteomics. Clin. Appl. 2008, 2, 556–570; doi:10.1002/prca.200780082.

40. Andersen, S.; Mischak, H.; Zürbig, P.; Parving, H.-H.; Rossing, P. Urinary Proteome Analysis Enables Assessment of Reno-protective Treatment in Type 2 Diabetic Patients with Microalbuminuria. BMC Nephrol. 2010, 11, 29; doi:10.1186/1471-2369-11-29.

41. Silva, S.; Bronze, M.R.; Figueira, M.E.; Siwy, J.; Mischak, H.; Combet, E.; Mullen, W. Impact of a 6-Wk Olive Oil Supplemen-tation in Healthy Adults on Urinary Proteomic Biomarkers of Coronary Artery Disease, Chronic Kidney Disease, and Diabetes (Types 1 and 2): A Randomized, Parallel, Controlled, Double-Blind Study. Am. J. Clin. Nutr. 2015, 101, 44–54; doi:10.3945/ajcn.114.094219.

42. Lohia, S.; Siwy, J.; Mavrogeorgis, E.; Eder, S.; Thoeni, S.; Mayer, G.; Mischak, H.; Vlahou, A.; Jankowski, V. Effect of Treat-ment with GLP-1R Agonists on the Urinary Peptidome of T2DM Patients. medRxiv 2023, 2023.05.03.23289439; doi:10.1101/2023.05.03.23289439.

43. Tye, S.C.; de Vries, S.T.; Wanner, C.; Denig, P.; Heerspink, H.J.L. Prediction of the Effects of Empagliflozin on Cardiovascular and Kidney Outcomes Based on Short-Term Changes in Multiple Risk Markers. Front. Pharmacol. 2021, 12, 786706, doi:10.3389/fphar.2021.786706.

44. Mukoyama, M.; Kuwabara, T. Role of Renin-Angiotensin System Blockade in Advanced CKD: To Use or Not to Use? Hypertens. Res. 2022, 45, 1072–1075; doi:10.1038/s41440-022-00902-7.

45. Heerspink, H.J.L.; Stefánsson, B. V; Correa-Rotter, R.; Chertow, G.M.; Greene, T.; Hou, F.-F.; Mann, J.F.E.; McMurray, J.J. V; Lindberg, M.; Rossing, P.; et al. Dapagliflozin in Patients with Chronic Kidney Disease. N. Engl. J. Med. 2020, 383, 1436–1446; doi:10.1056/NEJMoa2024816.

46. McMurray, J.J. V; Solomon, S.D.; Inzucchi, S.E.; Køber, L.; Kosiborod, M.N.; Martinez, F.A.; Ponikowski, P.; Sabatine, M.S.; Anand, I.S.; Bělohlávek, J.; et al. Dapagliflozin in Patients with Heart Failure and Reduced Ejection Fraction. N. Engl. J. Med. 2019, 381, 1995–2008; doi:10.1056/NEJMoa1911303.

47. Herrington, W.G.; Staplin, N.; Wanner, C.; Green, J.B.; Hauske, S.J.; Emberson, J.R.; Preiss, D.; Judge, P.; Mayne, K.J.; Ng, S.Y.A.; et al. Empagliflozin in Patients with Chronic Kidney Disease. N. Engl. J. Med. 2023, 388, 117–127; doi:10.1056/NEJMoa2204233.

48. Zinman, B.; Wanner, C.; Lachin, J.M.; Fitchett, D.; Bluhmki, E.; Hantel, S.; Mattheus, M.; Devins, T.; Johansen, O.E.; Woerle, H.J.; et al. Empagliflozin, Cardiovascular Outcomes, and Mortality in Type 2 Diabetes. N. Engl. J. Med. 2015, 373, 2117–2128; doi:10.1056/NEJMoa1504720.

49. Messerli, F.H.; Bangalore, S. Angiotensin Receptor Blockers Reduce Cardiovascular Events, Including the Risk of Myocardial Infarction. Circulation 2017, 135, 2085–2087; doi:10.1161/CIRCULATIONAHA.116.025950.

50. Hoang, V.; Alam, M.; Addison, D.; Macedo, F.; Virani, S.; Birnbaum, Y. Efficacy of Angiotensin-Converting Enzyme Inhibi-tors and Angiotensin-Receptor Blockers in Coronary Artery Disease without Heart Failure in the Modern Statin Era: A Meta-Analysis of Randomized-Controlled Trials. Cardiovasc. Drugs Ther. 2016, 30, 189–198; doi:10.1007/s10557-016-6652-7.

51. Tsujimoto, T.; Kajio, H. Spironolactone Use and Improved Outcomes in Patients with Heart Failure with Preserved Ejection Fraction with Resistant Hypertension. J. Am. Heart Assoc. 2020, 9; doi:10.1161/JAHA.120.018827.

52. Ferreira, J.P.; Cleland, J.G.F.; Girerd, N.; Pellicori, P.; Hazebroek, M.R.; Verdonschot, J.; Collier, T.J.; Petutschnigg, J.; Clark, A.L.; Staessen, J.A.; et al. Early and Late Renal Function Changes with Spironolactone in Patients at Risk of Developing Heart Failure: Findings from the HOMAGE Trial. Clin. Res. Cardiol. 2023, 112, 330–332; doi:10.1007/s00392-022-02116-w.

53. Mita, T.; Katakami, N.; Yoshii, H.; Onuma, T.; Kaneto, H.; Osonoi, T.; Shiraiwa, T.; Kosugi, K.; Umayahara, Y.; Yamamoto, T.; et al. Alogliptin, a Dipeptidyl Peptidase 4 Inhibitor, Prevents the Progression of Carotid Atherosclerosis in Patients with Type 2 Diabetes: The Study of Preventive Effects of Alogliptin on Diabetic Atherosclerosis (SPEAD-A). Diabetes Care 2016, 39, 139–148, doi:10.2337/dc15-0781.

54. Ishikawa, S.; Shimano, M.; Watarai, M.; Koyasu, M.; Uchikawa, T.; Ishii, H.; Inden, Y.; Takemoto, K.; Murohara, T. Impact of Sitagliptin on Carotid Intima-Media Thickness in Patients with Coronary Artery Disease and Impaired Glucose Tolerance or Mild Diabetes Mellitus. Am. J. Cardiol. 2014, 114, 384–388; doi:10.1016/j.amjcard.2014.04.050.

55. Chen, S.Y.; Kong, X.Q.; Zhang, K.F.; Luo, S.; Wang, F.; Zhang, J.J. DPP4 as a Potential Candidate in Cardiovascular Disease. J. Inflamm. Res. 2022, 15, 5457–5469; doi:10.2147/JIR.S380285.

56. Baksh, S.; Wen, J.; Mansour, O.; Chang, H.Y.; McAdams-DeMarco, M.; Segal, J.B.; Ehrhardt, S.; Alexander, G.C. Dipeptidyl Peptidase-4 Inhibitor Cardiovascular Safety in Patients with Type 2 Diabetes, with Cardiovascular and Renal Disease: A Retrospective Cohort Study. Sci. Rep. 2021, 11, 1–9; doi:10.1038/s41598-021-95687-z.

57. Xia, C.; Goud, A.; D’Souza, J.; Dahagam, C.H.; Rao, X.; Rajagopalan, S.; Zhong, J. DPP4 Inhibitors and Cardiovascular Out-comes: Safety on Heart Failure. Heart Fail. Rev. 2017, 22, 299–304; doi:10.1007/s10741-017-9617-4.

58. Yang, L.; Wu, X.; Wang, Y.; Wang, C.; Hu, R.; Wu, Y. Effects of Exercise Training on Proteinuria in Adult Patients with Chronic Kidney Disease: A Systematic Review and Meta-Analysis. BMC Nephrol. 2020, 21, 1–14; doi:10.1186/s12882-020-01816-7.

59. Winzer, E.B.; Woitek, F.; Linke, A. Physical Activity in the Prevention and Treatment of Coronary Artery Disease. J. Am. Heart Assoc. 2018, 7, 1–15; doi:10.1161/JAHA.117.007725.

60. Martínez-González, M.A.; Sayón-Orea, C.; Bullón-Vela, V.; Bes-Rastrollo, M.; Rodríguez-Artalejo, F.; Yusta-Boyo, M.J.; Gar-cía-Solano, M. Effect of Olive Oil Consumption on Cardiovascular Disease, Cancer, Type 2 Diabetes, and All-Cause Mortality: A Systematic Review and Meta-Analysis. Clin. Nutr. 2022, 41, 2659–2682; doi:10.1016/j.clnu.2022.10.001.

61. Marrone, G.; Urciuoli, S.; Di Lauro, M.; Ruzzolini, J.; Ieri, F.; Vignolini, P.; Di Daniele, F.; Guerriero, C.; Nediani, C.; Di Daniele, N.; et al. Extra Virgin Olive Oil and Cardiovascular Protection in Chronic Kidney Disease. Nutrients 2022, 14, 1–19; doi:10.3390/nu14204265.

62. Thompson, S.; Wiebe, N.; Gyenes, G.; Davies, R.; Radhakrishnan, J.; Graham, M. Physical Activity in Renal Disease (PAIRED) and the Effect on Hypertension: Study Protocol for a Randomized Controlled Trial. Trials 2019, 20, 1–10; doi:10.1186/s13063-019-3235-5.

63. Cattadori, G.; Segurini, C.; Picozzi, A.; Padeletti, L.; Anzà, C. Exercise and Heart Failure: An Update. ESC Hear. Fail. 2018, 5, 222–232; doi:10.1002/ehf2.12225.

